# Chronicity moderates the impact of severity on Central Executive - Default Mode Network functional interactions in Depression

**DOI:** 10.64898/2026.01.28.26345027

**Authors:** Tamires Zanao, Piergiorgio Salvan, Lais B. Razza, Pedro Henrique Rodrigues da Silva, Andre R Brunoni, Jacinta O’Shea

## Abstract

Neuroimaging has revealed that major depression is underpinned by dysfunctional brain networks, with symptom variability stemming from altered interactions within and between brain regions. While the effect of depression severity is well-studied, the effect of depression duration (chronicity) is relatively neglected, despite its clinical significance. This study examined how severity, chronicity, and their interaction affect brain network connectivity and grey matter volume. Forty-six patients (31 females, mean age 40.5) were assessed using whole-brain network modeling and voxel-based morphometry (VBM). Severity was measured via the Hamilton Depression Rating Scale, and chronicity was defined as an episode lasting over 24 months. The key finding was that chronicity moderated the impact of severity on functional connectivity between the Central Executive Network (CEN) and the precuneus (part of the Default Mode Network, DMN). Chronic versus non-chronic patients showed opposite patterns. Non-chronic patients showed stronger CEN-Default Mode Precuneus connectivity at low severity and weaker at high severity; chronic patients showed the reverse. This study reveals a novel impact of chronicity on CEN-DMN interactions, a neglected moderator of brain-symptom severity correlations in depression.

## Introduction

Major depressive disorder (MDD) is a widely prevalent and debilitating condition that is often resistant to treatment^1,2^. Core symptoms encompass depressed mood and anhedonia; however, clinical manifestations are markedly heterogeneous, with affective, cognitive, and somatic features variably represented across individuals^3^. The temporal progression of MDD, particularly its severity and chronicity, is a major contributor to the disease burden^4,5^. About 17% of MDD cases are moderate and 10% are severe ^6^. Chronic depression, typically defined as a current depressive episode lasting longer than 24 months, has a 4.6% lifetime prevalence and affects approximately 30% of those with a lifetime depressive disorder^7^. Both severity and chronicity play critical roles in the management of depression^5,8^, and are predictors of the outcome of short-term antidepressant treatment ^8,9^. Severe depression significantly impairs quality of life and daily functioning and may require more intense treatment, such as electroconvulsive therapy^10,11^. Chronic depression, in turn, is frequently even more incapacitating, as it is associated with greater suicidality, and poorer response to antidepressant treatments^12–15^. While severity and chronicity can co-occur and interact, severity has been more thoroughly investigated^11,16–18^, while chronicity has been relatively neglected ^19,20^.

Functional neuroimaging studies indicate that MDD is associated with dysregulated interactions among large-scale brain networks^21,22^,suggesting that disrupted intra- and inter-network connectivity contributes to symptom heterogeneity^23,24^. In parallel, meta-analytic findings from structural imaging report grey matter alterations in regions involved in emotion regulation and cognitive control, particularly within prefrontal and limbic circuits^25–28^. Structural abnormalities may be linked to these functional network changes ^29,30^. Together, structural and functional evidence offer distinct yet complementary perspectives on brain organization in MDD and may help elucidate how clinical features such as severity and chronicity relate to underlying neural mechanisms.

Grey matter alterations have been associated with both severity and chronicity^31–33^. While the impact of severity on functional network alterations has been relatively well studied, chronicity remains comparatively underexplored^34–36^. Fewer studies have included both functional and structural analyses in MDD^29^. Integrating structural and functional measures within the same framework therefore provides an opportunity to disentangle the distinct and overlapping neural signatures of severity and chronicity, offering a more comprehensive account of clinical heterogeneity in MDD.

In the present study, we adopt an exploratory, multimodal framework to examine how symptom severity, chronicity, and their interaction relate to both structural and functional brain organization in MDD^37–40^. Using baseline cross-sectional data from a clinical trial, we combined whole-brain Independent Component Analysis (ICA) of functional connectivity with voxel-based morphometry (VBM) of grey matter volume. Rather than restricting our analyses to specific regions or predefined networks, we aimed to characterize the broader landscape of neural variation associated with these key clinical dimensions. By examining structural and functional measures in parallel, we aimed to explore how severity and chronicity relate to differences in brain organization across modalities, thereby contributing to a more nuanced understanding of the neural correlates of severity and chronicity. We hypothesized that severity and chronicity represent at least partially dissociable constructs, reflected by distinct neural correlates, which would be more pronounced in patients who are both chronic and severe.

## Methods

### Study design

This study employed a cross-sectional design using baseline neuroimaging data obtained from participants enrolled in a clinical trial. Participants were patients diagnosed with MDD according to standard clinical and diagnostic criteria. Data collection occurred between October 2013 and July 2016 at the Institute of Psychiatry, Hospital das Clínicas, University of São Paulo Medical School, São Paulo, Brazil. The study protocol was reviewed and approved by the Research Ethics Committee of the Hospital das Clínicas, University of São Paulo Medical School (CEP-HCFMUSP) and by the National Research Ethics Commission (CONEP) of the Brazilian Ministry of Health. Ethical approval was granted under the Certificate of Presentation for Ethical Review (CAAE: 10173712.3.0000.0076). All participants provided written informed consent before enrolment.

### Patients

We included a subsample from the ELECT-TDCS study who underwent resting-state functional MRI and T1 structural (MRI) scans before the intervention. All patients with MDD (21-67 years old) were diagnosed according to the Diagnostic and Statistical Manual of Mental Disorders, fifth edition (DSM-5), by certified psychiatrists. Inclusion criteria comprised a score of ≥17 points on the Hamilton Depression Rating Scale (HDRS-17), low suicide risk, a minimum of 8 years of education to ensure understanding of informed consent, and willingness to adhere to the study protocol. Exclusion criteria included bipolar disorder, brain injury, pregnancy, specific contraindications to tDCS or MRI, current or previous use of escitalopram, and current or previous enrollment in tDCS trials. Anxiety disorder comorbidity was not excluded. Patients who were using antidepressants underwent a drug washout period before the study, with a minimum drug-free period of at least 5 drug half-lives. If benzodiazepines were used, they were permitted during the study at a maximum dosage of 20 mg per day (diazepam equivalent). The length of the current episode, longer than 24 months was considered chronic^41^. Severity was measured with the continuous score of the HDRS-17, with higher values meaning more severe symptomatology ^3,9,42^.

### MRI acquisition

Image acquisition and processing were conducted following standardized procedures before the first stimulation session. All images were acquired using a 3 T Philips Achieva MRI system equipped with an eight-channel coil at the Institute of Radiology within the Clinical Hospital of the University of São Paulo, São Paulo, Brazil. T1-weighted sequences were obtained utilizing a 3D FFE pulse sequence with the following parameters: field of view (FOV) of 240 × 240 × 180 mm^3^, spatial resolution of 1 × 1 × 1 mm^3^, repetition time (TR) of 7 ms, echo time (TE) of 3.2 ms, and flip angle (FA) of 80 degrees, resulting in 180 sagittal slices. For functional connectivity acquired in resting-state, the parameters included EPI single shot (FOV of 240 × 240×144 mm3, spatial resolution 3 × 3×4 mm3, TR = 2000 ms, TE = 30 ms, imaging matrix 80 × 79, FA = 80°, 32 slices, totalizing 200 volumes).

MRI was obtained for 68 out of the 245 patients enrolled in the original trial. The main reasons for missing MRI acquisition included MRI collection starting only after 1/3 of the sample had already been recruited; patient refusal, as MRI collection was optional, exclusion of patients due to MRI contraindications, and scheduling conflicts. Additionally, 19 MRI scans were excluded due to poor quality (head motion or abnormal anatomical findings), and 3 patients were excluded due to artifacts on resting-state images. This resulted in a total of 46 datasets being included. The demographic and clinical characteristics of these 46 patients are outlined in Table 1.

**Table 1.**
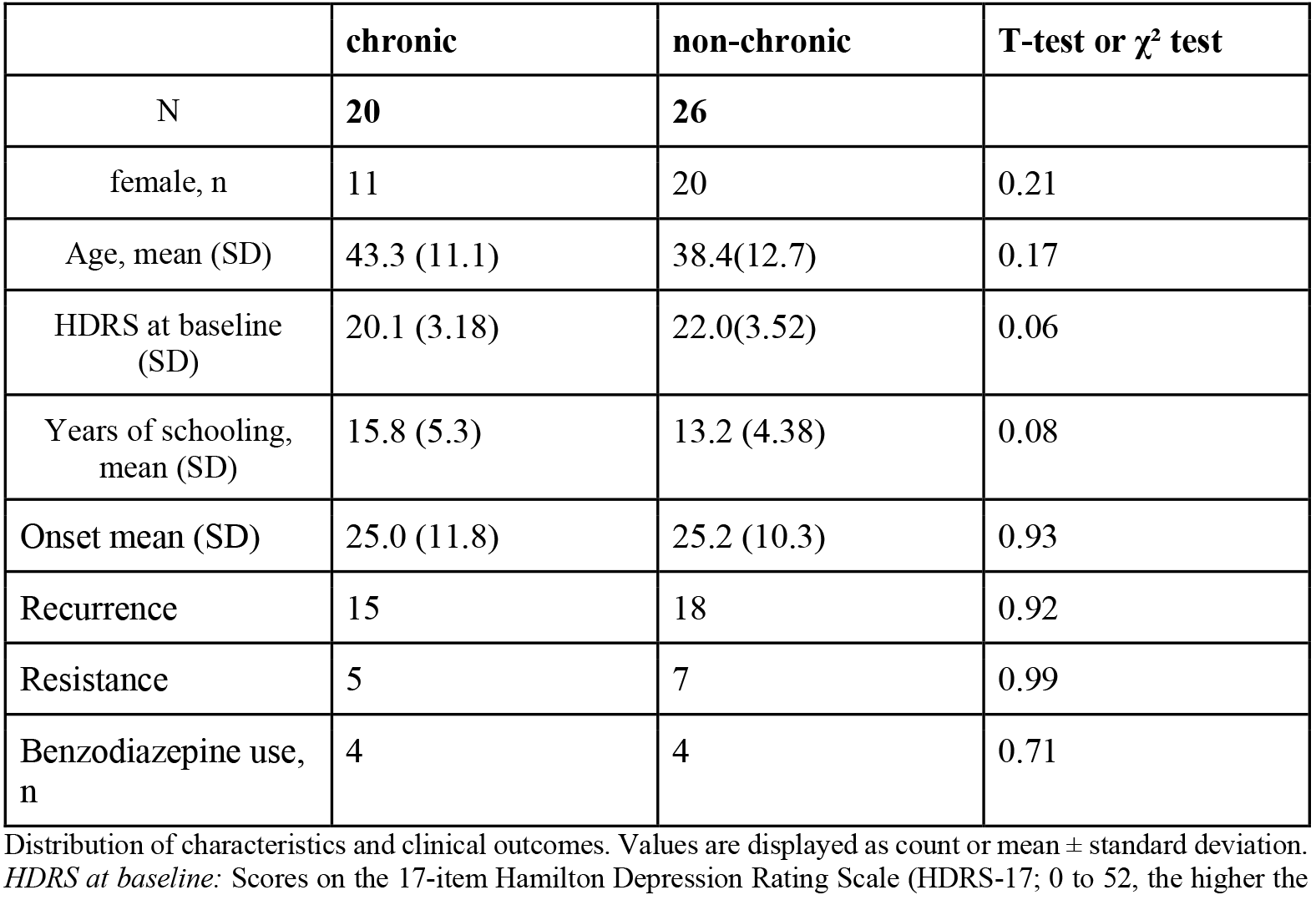
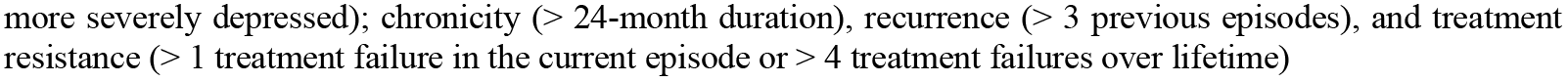
Patients’ characteristics.

|  | **chronic** | **non-chronic** | **T-test or $\chi^2$ test** |
| --- | --- | --- | --- |
| N | **20** | **26** |  |
| female, n | 11 | 20 | 0.21 |
| Age, mean (SD) | 43.3 (11.1) | 38.4(12.7) | 0.17 |
| HDRS at baseline (SD) | 20.1 (3.18) | 22.0(3.52) | 0.06 |
| Years of schooling, mean (SD) | 15.8 (5.3) | 13.2 (4.38) | 0.08 |
| Onset mean (SD) | 25.0 (11.8) | 25.2 (10.3) | 0.93 |
| Recurrence | 15 | 18 | 0.92 |
| Resistance | 5 | 7 | 0.99 |
| Benzodiazepine use, n | 4 | 4 | 0.71 |

Distribution of characteristics and clinical outcomes. Values are displayed as count or mean ± standard deviation. *HDRS at baseline*: Scores on the 17-item Hamilton Depression Rating Scale (HDRS-17; 0 to 52, the higher the
more severely depressed); chronicity ( $> 24$ -month duration), recurrence ( $> 3$ previous episodes), and treatment resistance ( $> 1$ treatment failure in the current episode or $> 4$ treatment failures over lifetime)

### MRI pre-processing

Brain extraction was performed in FSL, using Brain Extraction Tool (BET)^43^. Subsequently, FSL FAST ^44^ was used to tissue-type segmentation and to provide a bias-field corrected version of the brain-extracted structural images. Structural data were analysed with FSL-VBM (^45^ http://fsl.fmrib.ox.ac.uk/fsl/fslwiki/FSLVBM), an optimised VBM protocol^45,46^ carried out with FSL tools ^47^. First, structural images were brain-extracted and grey matter-segmented before being registered to the MNI 152 standard space using non-linear registration ^48^. The resulting images were averaged and flipped along the x-axis to create a left-right symmetric, study-specific grey matter template. Second, all native grey matter images were non-linearly registered to this study-specific template and “modulated” to correct for local expansion or contraction due to the non-linear component of the spatial transformation. The modulated grey matter images were then smoothed with an isotropic Gaussian kernel with a sigma of 5 mm. Finally, voxelwise GLM was applied using permutation-based non-parametric testing, correcting for multiple comparisons across space.

#### Registration

Rigid registrations between multimodal MRI native spaces were calculated using FSL FLIRT with a boundary-based cost function ^49,50^. Nonlinear transformations to the MNI152 standard-space T1 template were estimated through FSL FNIRT. Subsequently, we registered all the grey matter images to the template, modulated and smoothed them with different kernel sizes to reduce noise and preserve details, and ran GLM analysis using permutation testing. We performed randomization with 5000 permutations and sigma 5 for the Threshold-Free Cluster Enhancement (TFCE)^51^ based analysis. The nonlinear transformations were applied to the resting state (rs-fMRI) data.

#### Functional

Functional images were preprocessed using FSL (FSL 6.0.6. (https://www.fmrib.ox.ac.uk/fsl/index.html) and the Python-based re-implementation of the MATLAB-based <u>FSLNets</u> network modeling toolbox https://git.fmrib.ox.ac.uk/fsl/fslnets. All individual datasets were quality checked. Motion correction was carried out with MCFLIRT^49^, and co-registration between functional and structural images was visually inspected to ensure alignment quality, as head motion is known to strongly affect registration accuracy^52^. Motion-related noise was further addressed at the denoising stage: independent components identified as artifacts (e.g., edge, vascular, and motion-related patterns) were excluded based on established ICA classification criteria^53–55^. Temporal filtering was performed using a high-pass filter with a cutoff of 100 s (0.01 Hz) to remove low-frequency drifts while preserving the low-frequency fluctuations characteristic of resting-state fMRI (0.01–0.1 Hz; ^56,57^). Band-pass filtering was not applied, as the combination of high-pass filtering and ICA-based denoising has been shown to effectively reduce physiological noise while avoiding the removal of neural signal of interest ^58^. Finally, spatial smoothing was applied using a Gaussian kernel of 5 mm FWHM, approximately twice the voxel size, in line with the recommended practice of smoothing at 1.5–2 times voxel dimensions to improve signal-to-noise ratio while preserving anatomical specificity^59^. The rs-fMRI data were corrected for both intervolume and intravolume participant head motion and EPI distortions ^60^. Nuisance regression was performed to reduce physiological and non-neuronal noise, including signals extracted from white matter and cerebrospinal fluid (CSF). Independent components (IC) of single-participant data were manually classified to identify and remove artifacts, such as those caused by motion and physiological noise. The cleaned single-participant data were aligned to the standard space. In order to avoid bias and to provide comparison of our results with previous research, we used the UKbiobank group IC Analysis with 25-component (https://www.fmrib.ox.ac.uk/ukbiobank/). The set of spatial maps from the UK Biobank 25-component ICA was then used to generate participant-specific versions of the spatial maps and associated timeseries using dual regression ^61^. First, for each participant, the UK Biobank group-average spatial maps were regressed (as spatial regressors in a multiple regression) into the participant’s 4D fMRI dataset, producing a set of participant-specific timeseries, one per group-level spatial map. Next, these timeseries were regressed (as temporal regressors, in a multiple regression) back into the same 4D dataset, resulting in a set of participant-specific spatial maps, one per group-level map. We then tested for group differences using FSL’s randomise permutation-testing tool.

### Statistical analysis

Clinical and demographic statistics were conducted using R version 4.3.1.

#### Functional Network modeling

To investigate whether depression severity differently alters the interactions between large-scale brain network connectivity in chronic versus non-chronic MDD patients, we conducted functional network modeling using FSLNets (v0.6). This exploratory approach is well-suited for detecting variations in the strength of inter-network connectivity, which have been less frequently examined in this population. Resting-state fMRI data were preprocessed as described above, and group-level ICA was performed in MELODIC to identify spatially ICs. Dual regression was then applied: in the first stage, participant-specific time series were obtained by regressing the group ICA maps into each participant’s 4D data; in the second stage, these time series were regressed back into the data to produce participant-specific spatial maps, although this output was not used for connectivity modeling. We focused on the time series generated from stage one, which were visually inspected for quality and assessed via power spectra to identify noise components. From the ICA decomposition, 20 out of the 25 ICs extracted from the temporal concatenation at the UKbiobank^55^ were identified based on previous literature as resting-state networks of interest^62^. The ICs 3, 21, 22, 23, and 24 were identified as noise components (physiological or motion-related artifacts) based on ICA classification criteria, including high-frequency content and edge-related spatial patterns, and were excluded from further analysis^55^. FSLNets was used to estimate full correlation matrices representing functional connectivity between all pairs of retained components. Functional connectivity was estimated using full correlations between independent components, consistent with prior ICA-based network modeling approaches. Time series were regularized using the default ridge regression, which improves the reliability of correlation estimates by reducing the influence of noise and multicollinearity. A node represents an IC derived from group-level IC analysis, and an edge is the statistical association between the time series of two nodes. With 20 retained ICs, the analysis involved 190 unique undirected edges, defined as pairwise correlations between node (IC) time series. These edges represent the connectivity features entered into the second-level statistical modeling and index the strength of interactions between brain networks. Conceptually, they provide a means to characterize between-network connectivity implicated in depression and to assess the influence of symptom severity and chronicity on these interactions.

Group-level statistical analysis was performed using a GLM in the FSLNets GUI. Significance testing was conducted using TFCE applied in edge space to identify clusters of edges showing significant effects, controlling for FWE across all edges in the connectivity matrix. Unlike task-based fMRI analyses that use voxel-wise height thresholds and cluster-extent correction, network-based inference in FSLNets involves correction across the full set of edges derived from the connectivity matrix, allowing for comprehensive control of multiple comparisons. Specifically, the GLM included seven explanatory variables (EVs) to test whether depression severity had a differential impact on brain network connectivity in chronic vs. non-chronic MDD patients. The model was structured as follows:

EV1 coded group membership for non-chronic participants (1 for non-chronic, 0 for chronic),

EV2 coded group membership for chronic participants (1 for chronic, 0 for non-chronic),

EV3 included individual Hamilton Depression Rating Scale (HDRS) scores for the non-chronic group (values entered for participants coded as 1 in EV1, zeros otherwise),

EV4 included HDRS scores for the chronic group (values entered for participants coded as 1 in EV2, zeros otherwise),

EV5 modeled sex,

EV6 years of education, and EV7 age as covariates of no interest. Excluding EV1 and EV2, all other variables were demeaned across groups.

Two contrasts were specified to test the interaction between chronicity and depression severity: EV3 > EV4 (contrast weights: [0 0 1 −1 0 0 0]), which tested whether the association between HDRS and connectivity was stronger in non-chronic than chronic participants, and EV4 > EV3 (contrast weights: [0 0 −1 1 0 0 0]), which tested the opposite direction. This design allowed us to assess whether the relationship between depression severity and functional connectivity differed by chronicity group, controlling for sex, year of education and age. This enabled assessment of functional network connectivity and visualization of connectivity structure across subgroups. Edges showing significant interaction effects were extracted, and group-wise distributions were plotted to evaluate which network pairs contributed to the chronicity × severity interaction.

#### Functional connectivity correlations with grey matter

In the event of significant findings from the network modeling analysis, follow-up voxel-wise analyses were conducted to test for a relationship between functional connectivity and grey matter volume. For each participant, functional connectivity values for the significant functional connectivity edge were extracted from the FSLNets output using fslmaths and fslmeants, after identifying the relevant pair of independent components. These values corresponded to the Fisher z-transformed full correlation coefficients for that edge.

To test whether differences in functional connectivity were associated with differences in grey matter volume, we tested the following design matrix:

EV1 modeled the intercept (a column of ones), accounting for the overall mean grey matter volume across participants,

EV2 modeled the participant-specific functional connectivity values for the edge of interest, demeaned to center the predictor at zero.

EV3 demeaned modeled sex,

EV4 demeaned years of education, and

EV5 demeaned age as covariates of no interest.

Two contrasts were specified to test for positive ([0 1 0 0 0]) and negative ([0 –1 0 0 0]) correlations between functional connectivity and grey matter volume. Non-parametric permutation testing was performed with 5,000 permutations, and statistical significance was assessed using TFCE with FWE correction at p < 0.05.

#### Grey matter

Finally, we tested the unique effect of severity and chronicity on grey matter brain volume using FSL VBM. Following the BET, the grey matter partial volume images were then aligned to the MNI152 standard space using non-linear registration (FNIRT), modulated to correct for local expansion or contraction due to spatial normalization, and smoothed with an isotropic Gaussian smoothing kernel Full Width at Half Maximum (FWHM)(sigma = 5 mm). For the statistical analysis, a GLM was constructed using the randomise tool with 5,000 non-parametric permutations. To test for the positive effect of severity and chronicity on VBM differences, the design matrix included six EVs:

EV1 modeled the intercept (a column of ones, representing the mean grey matter volume across all participants),

EV2 modeled depression severity using individual HDRS scores, EV3 included age,

EV4 included years of education (scholarity),

EV5 coded chronicity (1 = chronic, 0 = non-chronic), and EV6 included sex.

All variables were demeaned. The contrasts were specified to test the unique effect of severity ([0 1 0 0 0 0]) and chronicity ([0 0 0 0 1 0]), while controlling for age, scholarity, and sex. In addition, an interaction term between severity and chronicity was included to evaluate potential combined effects. Family-wise error (FWE) correction was applied to all statistical tests.

## Results

### 1) Chronicity moderates the effect of severity on Central Executive Network x Default Mode Network Precuneus interactions

To test whether severity impacts differently on resting-state network interactions in chronic versus non-chronic depressed patients, we performed network modeling analysis. There was a positive correlation for the functional connectivity between IC 16 and IC 20 (p<0.05, adjusted for demeaned age, sex, and education). Following the nomenclature used by Lee et al. (2023) for the rs-fMRI ICs of the UK Biobank rs-fMRI ^63^ (figure 1A), from now on, IC 16 will be referred to as the Central Executive Network (CEN) and IC 20 as the Default Mode Network (DMN) Precuneus. The DMN was subdivided into three subnetworks in Lee’s classification, with IC 20 corresponding to the Default Mode Precuneus component. To visualize the relationship between depression severity and CEN-DMN Precuneus functional connectivity in the chronic versus non-chronic subgroups, Pearson’s correlation was performed. The chronic subgroup had a significant moderate-to-strong positive correlation (r = 0.65, R^2^= 0.42, 95%CI=0.29 to 0.84, df = 18, p = 0.002). The non-chronic subgroup had a moderate negative correlation (r=-0.504, R^2^=0.25, 95% CI= −0.75 to −0.15, df = 24, p=0.009) (figure 1B). In other words, in patients with chronic depression, CEN-DMN Precuneus connectivity was weaker in those with less severe symptoms and stronger in those with more severe symptoms. For those in the non-chronic group, CEN-DMN Precuneus connectivity was stronger in those with less severe symptoms and weaker in those with more severe symptoms. We also tested whether individual differences in functional connectivity between the CEN and the DMN Precuneus, the functional connectivity identified in the FSLNets analysis shown in Figure 1, a chronicity-by-severity interaction, correlated with grey matter volume. This analysis revealed a significant positive correlation between functional connectivity and grey matter volume, but the cluster did not survive correction, with only 4 voxels appearing (FWE-corr p<0.05; TFCE-corrected p = 0.042; 4 contiguous voxels; peak voxel at MNI coordinates x= −64, y= −48, z=10).

**Figure 1.**
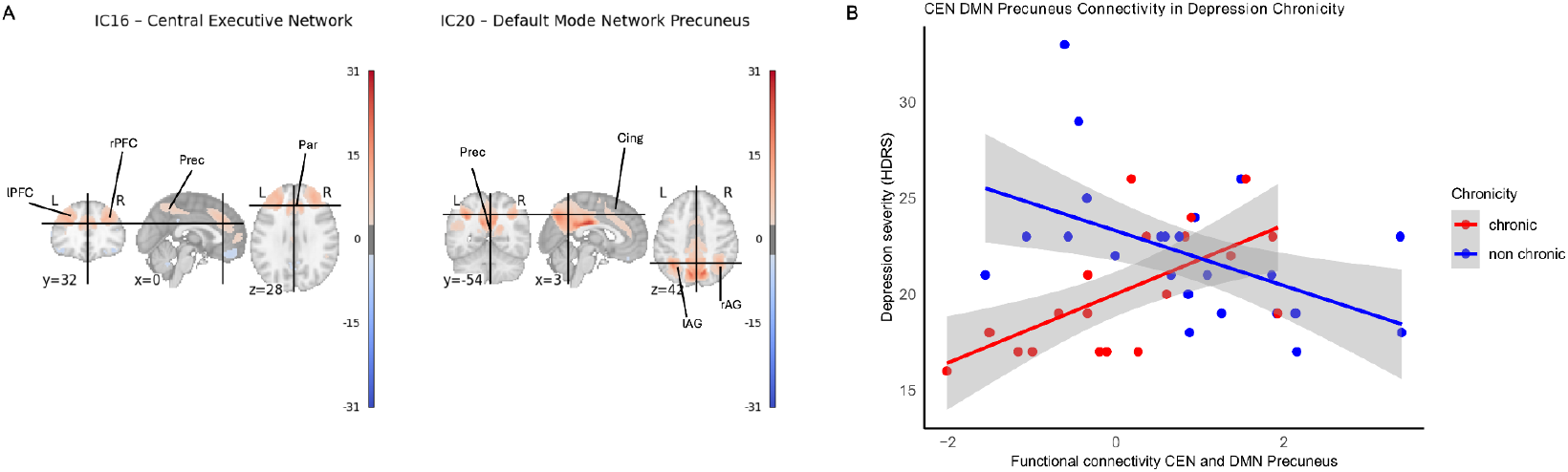
Chronicity moderates the effect of severity on CEN-DMN Precuneus functional connectivity. **A** Visualization of the IC 16 Central Executive Network (CEN) and IC 20 - Default Mode Network (DMN) Precuneus at MNI coordinates x=0, y=32 and z=28. Abbreviations: L, left; R, right; lPFC, left prefrontal cortex, rPFC, right prefrontal cortex; Prec, precuneus; Par, paracingulate gyrus; Cing, cingulate gyrus; lAG, left angular gyrus; rAG, right angular gyrus. **B** - **Opposite associations of CEN–DMN precuneus connectivity with depression severity in chronic vs. non-chronic patients**. Scatterplots show the relationship between functional connectivity of the CEN and the DMN Precuneus region and depression severity (HDRS scores). In chronic patients (red), greater CEN–DMN Precuneus connectivity was associated with higher severity, whereas in non-chronic patients (blue), greater connectivity was associated with lower severity. Shaded areas represent 95% confidence intervals. Chronic regression equation: y=20 + 1.8x, and R^2^=0.42; Non chronic regression equation: y= 23.3 - 1.43x and R^2^=0.25.

### 2) Positive correlation between severity and grey matter volume in two regions of the CEN (dorsal ACC and right DLPFC)

We tested the unique effect of severity and chronicity on grey matter brain volume using FSL VBM. We found a positive association with severity while adjusting for chronicity, age, sex and education (FWE-corr < 0.05) in the anterior cingulate/paracingulate gyrus(voxels = 351, peak voxel = 48, 79, 50, MNI coordinates =5, 31, 30, pmax value =0.004) and in the right dorsolateral prefrontal cortex (cluster 5, area 9/46D in the Sallet atlas, voxels = 31, peak voxel 37, 91, 55, pmax value = 0.953). Importantly, these clusters spatially overlapped with the network CEN (IC 16) identified in the functional connectivity network connectivity analysis (fig. 2). No significant result was found for chronicity while adjusting for severity, age, sex, and education (FWE-corr > 0.05). No significant interaction “severity-by-chronicity” was found (FWE-corr > 0.05). To assess the correlation between the HDRS scores and the grey matter for the chronic and non-chronic subgroups, Pearson’s correlation was performed. Both groups showed a significant positive correlation. In the chronic (standardized β = 0.545, SE = 0.198, 95% CI [0.135, 0.796], t(18) = 2.76, p = 0.013, R^2^= 0.297) and non chronic groups (β = 0.502, SE = 0.177, 95% CI [0.142, 0.744], t(24) = 2.84, p = 0.009, R^2^ = 0.252) showed greater GM volume significantly associated with higher depressive symptom severity, showing a moderate-to-strong positive effect.

**Figure 2.**
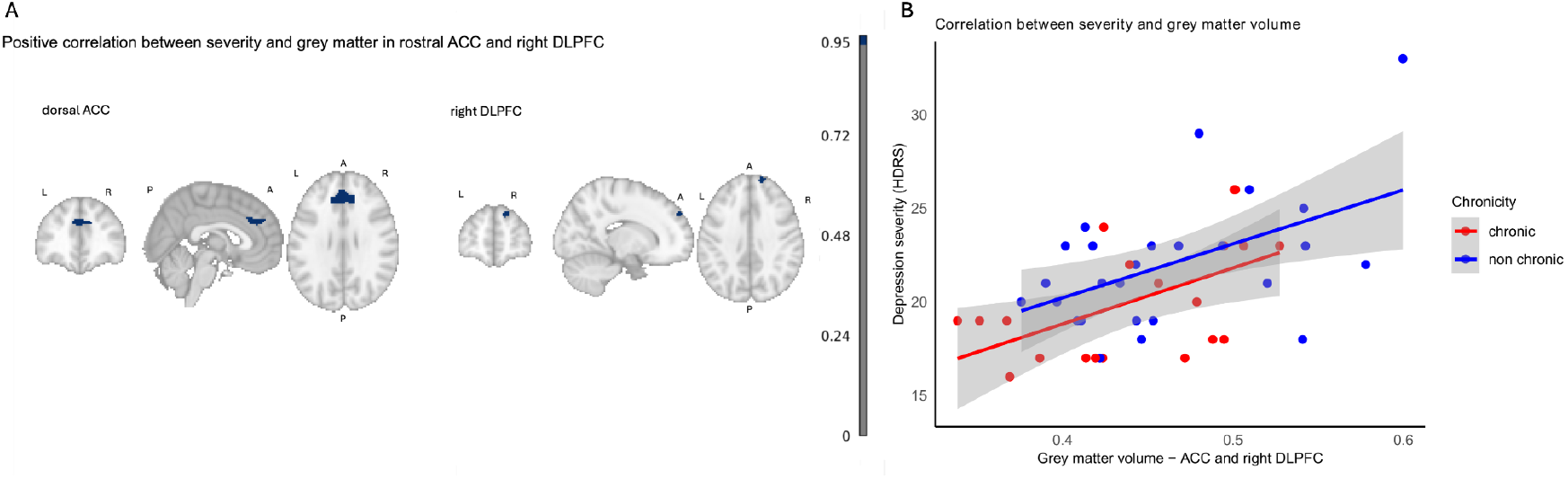
Positive correlation between grey matter volume and severity in dorsal ACC and right dorsolateral prefrontal cortex. **A** In blue, grey matter volume in anterior cingulate (ACC) and paracingulate cortex and right dorsolateral prefrontal cortex (DLPFC) positively correlated with depression severity (HDRS-17 scores). L, left; R, right; P, posterior, A, anterior. **B** Scatterplot shows individual patients’ grey matter volumes in the Anterior Cingulate Cortex (ACC) and right Dorsolateral Prefrontal Cortex (DLPFC) shown in panel A, and their association with higher depression severity for both chronic and non-chronic groups. Shaded areas represent 95% confidence intervals. Chronic regression equation: y=6.84 + 30 x, and R^2^=0.30; Non chronic regression equation: y= 8.69 + 28.8 x and R^2^=0.25.

## Discussion

In this exploratory study, we investigated how depression severity and chronicity, as well as their interaction, relate to brain organization by integrating functional connectivity and structural morphometry. At the functional level, we observed opposing severity-related patterns of CEN–DMN Precuneus connectivity depending on chronicity: in non-chronic patients, greater severity was associated with reduced connectivity, whereas in chronic patients, greater severity was associated with increased connectivity. This crossover effect indicates that the impact of symptom burden on network interactions may vary according to illness course, underscoring the importance of considering chronicity when interpreting functional alterations in MDD. At the structural level, depression severity—but not chronicity—was f associated with grey matter volume, with larger dorsal ACC and right DLPFC volumes associated with more severe symptoms. Of note, the ACC mainly overlapped with the CEN.

Understanding the significance of our findings requires consideration of the roles of the CEN and DMN, two large-scale networks that are central to cognitive control and internally oriented thought, respectively. The CEN, with key nodes in the dorsolateral prefrontal cortex and posterior parietal cortex, is primarily engaged during task-oriented activities requiring executive control^64^. In contrast, the DMN is a distributed network encompassing the medial prefrontal cortex, medial parietal cortex, angular gyrus, precuneus, posterior cingulate, and posterior hippocampus ^65^. It typically deactivates during externally focused tasks and engages during internally directed processes such as self-reflection, autobiographical memory, prospection, mind-wandering, and social cognition. During rest and quiet wakefulness, the DMN is thought to integrate past experiences with present feelings and future expectations ^66^. Given its complexity, the DMN is often divided into subnetworks, including limbic and precuneus divisions^63^.

The CEN and DMN are generally considered functionally opposing systems, with their interactions mediated by the salience network (SN). In healthy individuals, the SN flexibly regulates switching between externally focused CEN activity and internally directed DMN activity in response to environmental demands^64^. According to the Triple Network Model, depression reflects a disruption of this balance, with dysfunctional interactions among the DMN, CEN, and SN^22,67^. Consistent evidence of DMN hyperconnectivity and CEN hypoconnectivity has been linked to core clinical features of MDD, including rumination^68^ and negative bias^34^. A meta-analysis of resting-state connectivity studies suggested that altered connectivity between executive control systems and circuits supporting internal attention and emotional processing represent common neuroimaging findings in depression^34^.

The precuneus is a functional core of the DMN^69^, yet recent evidence highlights its broader role as a hub of large-scale brain networks, with strong involvement not only in the DMN but also in executive control systems^70^. Within the DMN, anterior and posterior subdivisions of the precuneus have been distinguished: the anterior portion is linked to theory of mind, and self-referential processes, whereas the posterior portion is more strongly related to episodic memory and visuo-spatial imagery ^70,71^. Finer-grained network parcellations, such as the Yeo–Buckner 17-network model, further suggest that the precuneus also interfaces with a CEN-related subnetwork (sometimes referred to as a para-cingulate network) positioned near the posterior cingulate and DMN core^62^. Evidence from parcellation, graph theoretical, and independent component analysis converges to support the precuneus as a structurally and functionally heterogeneous region bridging the DMN and CEN^62727374^.

The UKBiobank studies^55^ demonstrated that, in healthy individuals, the CEN and the DMN Precuneus were positively correlated. Our study did not include a healthy control group, however, non-chronic patients in our sample showed connectivity patterns more similar to those reported in healthy individuals, particularly among those with lower depression scores. Evidence supports that impairment of intrinsic functional connectivity throughout the course of depression is a progressive process^13^. Liu et al^13^ showed that while in the first episode MDD patients showed hypoconnectivity in different networks, hyperconnectivity was reported for others, including in the CEN and DMN, for recurrent depression. In our study, we observed a similar pattern in participants with chronic depression, particularly for the CEN–DMN Precuneus functional connectivity. Notably, a high percentage of our sample have also suffered recurrent depression, and recurrence and chronicity may co-occur.

In structural analyses only depression severity was associated with grey matter volume, for both ACC and right DLPFC. Both regions’ involvement in depression has been extensively explored ^25,27^, and their association with severity has been reported previously ^12,37,75^. ACC anatomically connects with dorsal neocortical and paralimbic areas, functionally acting in mood regulation and cognitive processing ^76,77^. Although higher volume is often interpreted as an indicator of enhanced cognitive functioning ^46^, and previous meta-analyses have reported grey matter reductions in ACC in MDD ^27,26,78^, other studies have found contrast findings. For example, a meta-analysis^79^ of VBM studies suggested that reductions in ACC volumes were associated with antidepressant intake at the time of scanning. Therefore, studies of unmedicated patients, such as ours, could perhaps better elucidate brain abnormalities directly related to MDD itself. As in our study, Zhao et al.^79^ reported increased grey matter in bilateral ACC in medication washed-out patients. Another study indicated lifetime MDD (experienced at least one episode of MDD at any point of their lives) was associated with higher ACC volume in women^80^, which comprises almost 70% of our sample. The DLPFC has been a focus of neuroimaging studies in MDD ^81^. More than a key component of the CEN involved in prefrontal-limbic circuitry, dysregulation in the DLPFC is linked to poor emotional and cognitive control in MDD^81^. Positron Emission Tomography (PET) and repetitive Transcranial Magnetic Stimulation (rTMS) findings support the imbalance hypothesis of MDD, suggesting prefrontal asymmetry underlying symptoms with hypoactivity in the left DLPFC and hyperactivity in the right DLPFC ^75,81,82^. The valence-lateralization theory further posits that the (hypo-active) left prefrontal cortex processes positive emotions, while the (hyper-active) right processes negative emotions^83^. Our finding of increased right DLPFC volume is consistent with this framework.

This study has several limitations. The sample size is relatively small (N = 46; chronic n = 20, non-chronic n = 26). This limits the generalizability of our findings. It also means that null results (eg: grey matter x chronicity) may simply reflect insufficient power. Potential confounding factors related to both severity and chronicity (such as early onset, lifetime depression, treatment resistance, use of benzodiazepine, and clinical recurrence) may have influenced our results. However, we opted not to include an extensive set of covariates in the GLM analyses, as this could introduce methodological issues including overfitting, multicollinearity, and diminished statistical power. Although we defined severity using the HDRS, a well-validated and widely used scale for assessing MDD symptomatology, and operationalized chronicity according to DSM-5 criteria, residual confounding cannot be excluded. Third, the inclusion criterion of an HDRS score greater than 17 resulted in the selection of patients with moderate to severe depression, which may limit the applicability of our findings to individuals with milder forms of MDD. Fourth, whole-brain grey matter volume was not included as a covariate in the connectivity analyses. This decision was made to avoid obscuring region-specific structure–function associations and to maintain comparability with prior ICA-based connectivity studies. Nonetheless, this approach leaves open the possibility that global structural differences may have contributed to the observed effects. Finally, the relatively short duration of resting-state fMRI acquisitions may introduce moderate-to-low test–retest reliability^84^, potentially affecting the reproducibility of our results. Future research should consider strategies to improve reliability, such as increasing scan duration or combining multimodal imaging approaches. Despite these limitations, the inclusion of MDD patients in antidepressant washout represents a key strength of this study, allowing the assessment of brain alterations associated with the disorder itself rather than medication effects.

To the best of our knowledge, this study represents one of the first efforts to characterize inter-individual alterations in functional connectivity associated with both the severity and chronicity of MDD, in conjunction with grey matter analyses. Future investigations should aim to address the aforementioned limitations by recruiting larger and more representative samples to enhance statistical power and generalizability. Moreover, the integration of complementary neuroimaging modalities, such as diffusion tensor imaging (DTI), may yield a more comprehensive and multidimensional understanding of the neural substrates underlying chronicity and severity of MDD. From a clinical perspective, the present findings contribute to the growing effort to bridge neuroimaging research and psychiatric practice. By demonstrating distinct patterns of CEN–DMN Precuneus connectivity associated with depression severity and chronicity, our results underscore the importance of considering illness courses when interpreting neural alterations in MDD. This differentiation may help explain the heterogeneity of clinical presentations and treatment responses observed in depressive disorders. Although the current findings are preliminary and derived from a cross-sectional design, they provide a foundation for future longitudinal and treatment studies aimed at identifying neural markers predictive of chronicity, treatment resistance, or recovery trajectories in MDD.

In conclusion, this exploratory study provides novel insights into the interplay between depression severity, chronicity, and large-scale brain network organization in MDD. Our findings reinforce the involvement of the DMN and CEN,core components of the Triple Network Model, in the neurobiology of depression. These results suggest that severity and chronicity may represent distinct but interacting dimensions of MDD, each associated with specific patterns of network dysfunction. The observed differential effects of severity on CEN and DMN connectivity between chronic and non-chronic patients may help explain individual differences in symptom persistence and treatment responsiveness. Furthermore, the association between grey matter volume in the dorsal ACC and right DLPFC with severity highlights potential neuroanatomical correlates of cognitive and emotional regulation deficits commonly observed in clinical practice. Together, these findings advance our understanding of the neural substrates of MDD and may inform future psychiatric research aimed at improving diagnostic stratification, prognosis, and treatment personalization.

## Author contributions

TZ, PS, ARB, JOS: conceptualization, data curation, formal analysis, investigation, methodology, software, validation, visualization, writing—original draft preparation, writing—review and editing. LBR, PHRS: methodology, data acquisition, writing—review and editing.

## Data availability statement

Owing to the ethically sensitive nature of the research, supporting data cannot be made openly available. Anonymized data can be provided on request.

## Copyright

For the purpose of open access, the author has applied a CC BY public copyright license to any Author Accepted Manuscript version arising from this submission.

